# A novel, scenario-based approach to comparing non-pharmaceutical intervention strategies across nations

**DOI:** 10.1101/2023.09.14.23294544

**Authors:** Justin M. Calabrese, Lennart Schüler, Xiaoming Fu, Erik Gawel, Heinrich Zozmann, Jan Bumberger, Martin Quaas, Gerome Wolf, Sabine Attinger

**Affiliations:** Center for Advanced Systems Understanding (CASUS), Untermarkt 20, 02826 Görlitz; Helmholtz-Zentrum Dresden-Rossendorf (HZDR), Bautzner Landstraße 400, 01328 Dresden; Dept. of Ecological Modelling, UFZ - Helmholtz Centre for Environmental Research, Leipzig, Germany; Dept. of Biology, University of Maryland, College Park, MD, USA; Research Data Management - RDM, UFZ – Helmholtz Centre for Environmental Research, Leipzig, Germany; Dept. Monitoring and Exploration Technologies, UFZ – Helmholtz Centre for Environmental Research, Leipzig, Germany; Dept. of Economics, UFZ – Helmholtz Centre for Environmental Research, Leipzig, Germany; Leipzig University, Institute for Infrastructure and Resources Management, Leipzig, Germany; German Centre for Integrative Biodiversity Research (iDiv) Halle-Jena-Leipzig, Germany; ifo Institute - Leibniz Institute for Economic Research, Munich, Germany; Dept. of Computational Hydrosystems, UFZ - Helmholtz Centre for Environmental Research, Leipzig, Germany

**Keywords:** COVID-19, NPI, modelling, epidemiological, behavioural, macroeconomic

## Abstract

Comparing COVID-19 non-pharmaceutical intervention (NPI) strategies across nations is a key step in preparing for future pandemics. Conventional comparisons, which rank individual NPI effects, are limited by: 1) vastly different political, economic, and social conditions among nations, 2) NPIs typically being applied as packages of interventions, and 3) an exclusive focus on epidemiological outcomes of interventions. Here, we develop a coupled epidemiological-behavioural-macroeconomic model that allows us to transfer NPI strategies from a reference nation to a focal nation while preserving the packaged nature of NPIs, controlling for differences among nations, and quantifying epidemiological, behavioural and economic outcomes. As a demonstration, we take Germany as our focal nation during Spring 2020, and New Zealand and Switzerland as reference nations with contrasting NPI strategies. We show that, while New Zealand’s more aggressive strategy would have yielded modest epidemiological gains in Germany, it would have resulted in substantially higher economic costs while dramatically reducing social contacts. In contrast, Switzerland’s more lenient NPI strategy would have prolonged the first wave in Germany, but would have also have increased relative costs. Our results demonstrate that Germany’s intermediate strategy was effective in quelling the first wave while mitigating both economic and social costs.

## Introduction

The COVID-19 pandemic caught the world unprepared and exposed critical weaknesses in national pandemic response plans. Within four months of its emergence in Wuhan, China, the disease had spread globally, and the World Health Organisation (WHO) upgraded COVID-19 to pandemic status in March 2020. Most national governments were not willing to allow unchecked spread of SARS-CoV-2, but effective countermeasures were challenging and costly to implement. Non-pharmaceutical interventions (NPIs), such as mask mandates, lockdowns, and school closures, quickly became essential tools for combating the incipient COVID-19 pandemic [1–4]. NPIs were, consequently, applied around the world, but in heterogeneous ways and with diverse outcomes. Even though the pandemic has not yet come to a complete standstill worldwide (e.g., China), we believe that now is the right time to make retrospective comparisons among the contrasting NPI approaches employed by different nations to identify effective intervention strategies, and to be better prepared for future pandemics.

Multiple studies have assessed NPI deployment across nations and have attempted to rank and evaluate NPIs in terms of their average epidemiological effectiveness [1, 5–8]. Most commonly, NPIs are understood as effective if they impact specific epidemiological indicators, in particular if they reduce the basic reproduction number, *R*_0_, or COVID-19-related mortality [9–12]. Furthermore, several studies have compared how NPI effectiveness varied among nations (Bo et al. [9], Liu et al. [10]), relying on data sets such as the Johns Hopkins Coronavirus Resource Center [13] or the Oxford COVID-19 Government Response Tracker [14]. Other studies have assessed NPI impacts in selected countries [15], on a regional scale [11, 16], or examined effectiveness in individual countries [17, 18]. Overall, NPIs such as school closures, restricting gatherings, banning public events or mandating masks were found to be effective by a range of studies [10, 19].

While providing a useful baseline, such comparisons are difficult to perform and subject to a number of complications. First, NPI comparisons typically seek to isolate the effects of individual NPIs, but there are very few situations in which individual measures have been applied in isolation. Instead, interventions tend to be applied as packages, where the package composition, the application sequence, and the timing of application are all important (e.g., [20–23]). Second, cross-national comparisons are frequently confounded by each country having its own set of circumstances including governance, financial, and public health systems as well as education levels, political attitudes, income distributions, and myriad other factors, which we collectively refer to as National Framework Conditions (NFCs) [24]. Finally, NPI effectiveness has primarily been assessed from an epidemiological perspective, despite widespread evidence of marked variation in social and economic consequences of such interventions. A more balanced approach would also account for social and economic outcomes when accessing NPI performance (e.g., [15, 25]; for sustainability implications see [26]).

Here, we propose a novel, scenario-based method of comparing NPI effectiveness across nations that treats NPIs as packages of interventions, can control for NFCs, and integrates economic and social considerations into the assessment. Our approach is based on an SIR-type disease model coupled to elemental behavioural [27] and economic models [28], and can be fit to data from different nations. Following Schüler et al. (2021) [29], we estimate the time-dependent disease transmission rate for each nation as a piece-wise constant function with breakpoints defined by the times of NPI implementation or removal. By treating bundles of interventions as a package impacting the overall time-dependent transmission rate, this approach obviates the need to identify separate effects of individual interventions and shifts the focus to intervention strategies. It is then possible to quantify how a focal nation’s epidemiological, social, and economic outcomes would have changed under the NPI strategy of a reference nation via “what-if” scenario simulation. Specifically, we transfer a relativised version of the time-dependent transmission rate estimated from the reference nation into the focal nation’s model, simulate the epidemiological dynamics forward in time, and then calculate the resulting social and economic costs.

For simplicity, we focus our case study on the first COVID-19 wave in Spring 2020, well before vaccines became widely available. Additionally, we take Germany (DE) as our focal nation, and we consider scenarios from reference countries that employed more stringent (New Zealand, NZ) and less stringent (Switzerland, CH) NPI strategies relative to Germany. We find that the New Zealand scenario would have more quickly brought the first COVID wave in Germany under control, but at the expense of much larger reductions in social contacts than either the CH or DE scenarios. Additionally, compared to the strategy DE actually implemented, both the CH and NZ scenarios would have resulted in substantially higher economic costs, but for contrasting reasons. Finally, we demonstrate starkly different epidemiological outcomes based on differential social dynamics between countries, particularly when transferring the CH scenario to DE.

## Materials and methods

### Background: COVID-19 response in Germany, New Zealand, and Switzerland during spring 2020

Many NFCs are correlated with COVID-19 outcomes across nations. Specifically, the age structure, obesity, urbanisation, GDP *per capita*, and trust in the government are NFCs with higher correlation coefficients [30–33]. It is therefore critical to control for variation in these important NFCs in cross-national comparisons, which we do here in two ways. First, we have chosen reference countries that are in many ways similar to Germany. Second, we transfer the relativised contact rates from the two reference countries to the focal country, which we describe in detail in the Scenario Transfer section below.

Comparing key NFCs across our case study nations, we see that these three countries are quite similar in terms of age structure, obesity, urbanization, and the trust in their governments compared to world averages (Fig 1).

**Fig 1.**
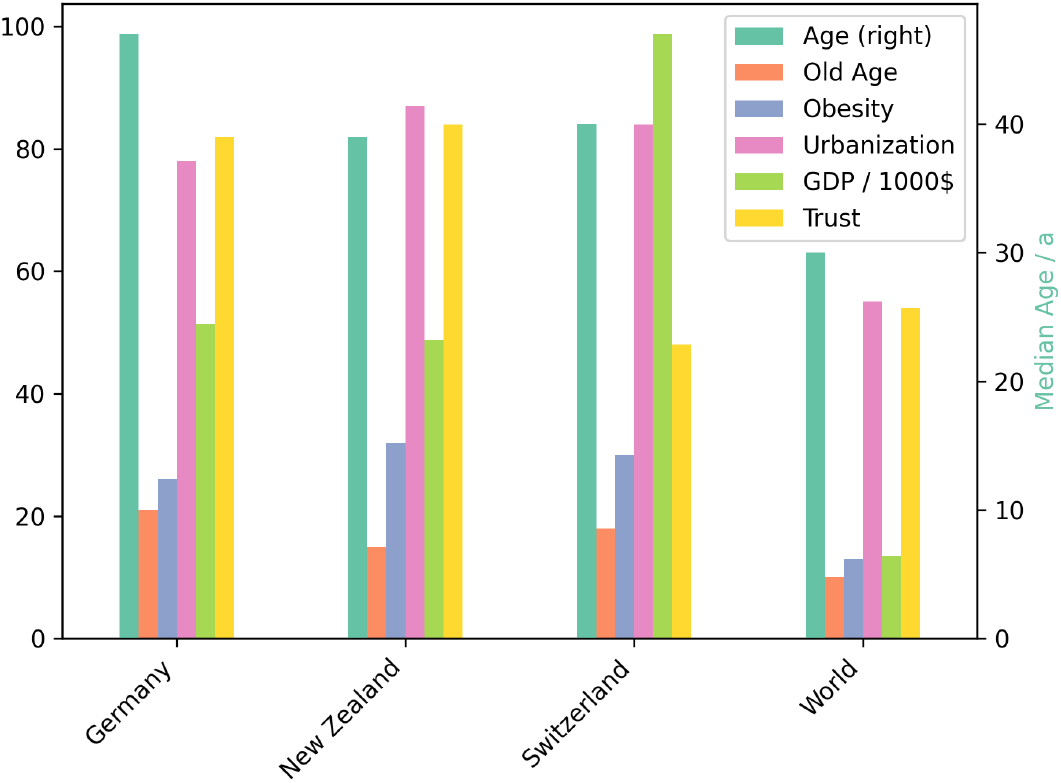
A comparison of key NFCs between the focal countries, the reference country and the world. The NFCs are the median age (Age, with the respective y-axis on the right), the percentage of people being 65 years or older (Old Age), the percentage of people having a BMI of over 30 (obesity), the percentage of people living in an urban area (Urbanisation), the nominal GDP per capita in 1000$, and the percentage of people trusting their government (Trust) [34].

We identified key milestones and turning points in the national responses to the COVID-19 pandemic based in early 2020 primarily on the COVID-19 Government Response Tracker data set [14] and supplemented with sources specific to each nation [35–41]. Figure 2 shows, for each of the three nations, the 7d incidence against the stringency index, which is a composite indicator tracking nine NPIs on a scale between 0 and 100 [14]. In Annex A, we break down the index into its nine constituents. A particular focus for comparing different response strategies is placed on the early days of the pandemic, as the reproduction number of the original variant of COVID-19 without intervention was estimated to exceed 1 in most cases [42, 43]. Thus, not only the intensity of NPIs is relevant, but the timing of their deployment is crucial.

**Fig 2.**
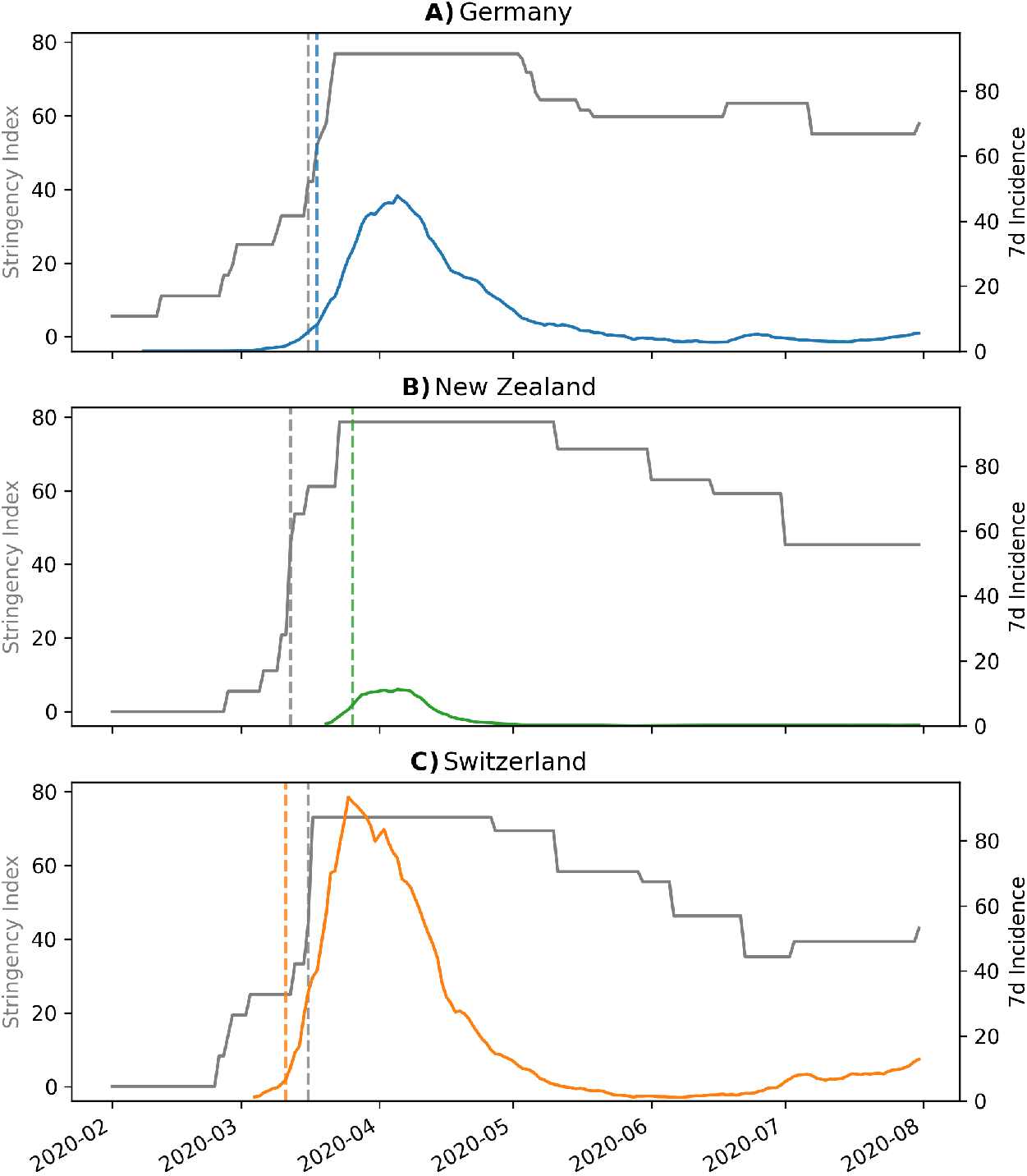
A comparison of the development over time of the stringency index (grey, left y-axis) and the 7d incidence (right y-axis) for A) Germany, B) New Zealand, and C) Switzerland. The grey vertical lines indicate the day at which the stringency index surpasses the threshold of 40. The coloured vertical lines indicate the day at which the time derivative of the 7d incidence surpasses the threshold of 1.5 d^*−*2^.

Germany implemented NPIs in a step-wise manner (Fig. 2A), including the cancellation of large-scale events on March 10, the closing of schools and kindergartens in most federal states by March 16, and culminating in a set of NPIs restricting private meetings and public life on March 22 [35]. During the same time, NZ (Fig. 2B), and CH (Fig. 2C) adopted a more stringent and a more lenient response, respectively. In CH, NPIs were deployed comparatively late, initially relying on public information campaigns and the restriction of large gatherings. Reacting to the strong increases in reported case numbers (Fig. 2C), CH implemented a variety of NPIs effective immediately on March 16-17, including the closure of schools, non-essential workplaces and the banning of public events [44]. In contrast, NZ issued quarantine orders for international arrivals by March 14, banned indoor gatherings of more than 100 individuals by March 19, and enacted a detailed four-tiered pandemic plan by March 21, 2020 at comparatively low levels of infection. By March 25, the highest alert level (‘Lockdown’) was announced, which banned all gatherings and travel, while mandating a strict stay-at-home order [40].

To account for differential timing of SARS-CoV-2 arrival, we now compare the timing of NPI implementation relative to the timing of fast growth in case counts across the three example nations. For each country, we therefore set a stringency index threshold of 40 and an incidence growth rate threshold of 1.5 d^*−*2^. Comparing the time lags between dates at which these thresholds where crossed, we see that NZ implemented strict NPIs 14 days before reaching the incidence growth threshold, while DE began NPI implementation only 2 days before crossing the growth threshold. In contrast, CH was slower to react and began NPI implementation in earnest 4 days *after* the growth threshold was exceeded. Though our chosen thresholds are somewhat arbitrary, the pattern of time lags across nations is robust to the choice of thresholds.

Following reductions in reported infection levels throughout the following weeks, all three nations responded with changes in NPI deployment. Germany and CH both gradually eased NPI stringency, albeit with differences in timing and the extent to which restrictions were lifted (cf. Fig. 2A, C): CH re-opened schools and non-essential commercial establishments earlier and permitted contacts and public events under fewer restrictions. In NZ, NPIs were adjusted based on the pandemic plan, reducing NPI stringency in three steps as the reported infection levels fell (Fig. 2B).

### Coupled model analysis

Here, we define each component of our coupled epidemiological-behavioural-macroeconomic model, specify how the components interact, and then describe how we transfer estimated rate functions from reference countries to DE. We specifically aim to keep the complexity of our models at a minimum to preserve parameter identifiability, which allows us to directly link all model components to data.

#### Epidemiological-behavioural model

We begin with a simple SIR compartment model to represent the epidemiological dynamics

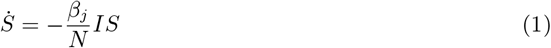

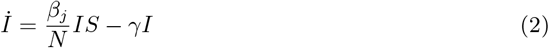

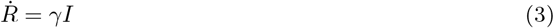

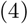

where *S, I*, and *R* are the number of susceptible, infected, and recovered individuals, and *N* = *S* + *I* + *R*. The parameters of this model are the recovery rate *γ* and the piece-wise contact rate *β*_*j*_, where *j* indexes the dates at which national NPIs are introduced or lifted [29].

To incorporate behaviour in our model, we note that an individual’s perceived risk of an infection, personal risk aversion, valuation of social contacts, and potential for income loss may affect their decisions [45]. The resulting autonomous behavioural adjustments that individuals make in response to an outbreak, when averaged over the population, can substantially affect epidemiological dynamics [27]. Population-level differences in behavioural responses may therefore represent an important source of variation in COVID outcomes among nations. To capture this potential variation in our coupled model, we assume that individuals derive utility *u*(*β*_*r*_ ) from social contacts, and following [27], model utility as

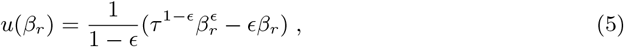

with *β*_*r*_ *ϵ* (0, 1] being the relative contacts with respect to the pre-pandemic contact level and *ϵ ϵ* (0, 1) captures how important it is to enjoy at least some contacts. The variable *τ ϵ* (0, 1] models the contact reduction that results from the NPIs. If there are no NPIs, we set *τ* = 1 and the stricter the NPIs, the lower the value of *τ*. The level of social contacts that maximises the utility is 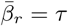, and the corresponding level of utility is normalised to 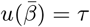. In an epidemic situation where almost all individuals are susceptible, the individual risk of an infection can be approximated by *βIS/N ≈ βI*.

A rational, risk-averse individual chooses their contacts as a trade-off between utility from contacts and the expected utility loss from an infection Δ*v* (see Annex B). The number of contacts is determined by

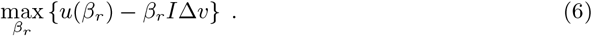

Solving this yields

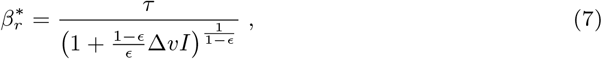

and with 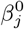 being the contact rates without the behavioural model, we can calculate the contact rates for the SIR model (1) - (3) with 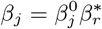. Following [27], we set *ϵ* = 0.7. The parameter *τ* depends on the stringency of the current NPIs, and Δ*v* depends on individual risk assessment and individual risk aversion and we assume it to be constant over the duration of our scenarios.

The recovery rate *γ* of the epidemiological model, the stringency index *τ*, and the risk aversion Δ*v* of the behavioural model are estimated inversely by solving a nonlinear least-squares problem.

#### Macroeconomic model

The scenario analysis for economic activity is based on a model developed in [28]. First, we model economic activity as a function of the time-dependent reproduction number, *R*_*t*_, following empirical relationships at the industry level to account for heterogeneous effects of an aggregate NPI policy. Second, we simulate economic activity associated with different NPI scenarios. A key assumption in this analysis is that the reproduction rate *R*_*t*_ is proportional to the aggregate NPI policy such that *R*_*t*_ is lower the stricter the interventions.

To model economic activity as a function of *R*_*t*_, we measure economic activity over two time windows and link it to the associated reproduction numbers at those times. The first time window, *t*_1_, is immediately after the first lockdown began to be lifted. The second time window, *t*_2_, is after several step-wise relaxations of policy measures. We estimate the associated reproduction numbers as follows. First, we fit the SIR model (1) - (3) to the entire case data timeseries. Then, for each day in a given time window (*t*_1_ or *t*_2_), we extract the model-fitted value of case numbers and estimate *R*_*t*_ according to Cori et al. 2013 [46]. Finally, we average *R*_*t*_ values over all days in a given time window. Repeating this procedure separately for *t*_1_ and *t*_2_ yields the estimates *R*_*t*1_ and *R*_*t*2_. Finally, we regress measures of monthly economic activity *y*_*m*_(*t*_1_) and *y*_*m*_(*t*_2_) against *R*_*t*1_ and *R*_*t*2_ to estimate a linear relationship between economic activity and the reproduction number at the industry level.

Specifically, the German government announced partial re-openings on April 20, so we set the first time window to be April 27 to May 2, 2020 and estimated *R*_*t*1_ = 0.84 (for comparison, the official RKI nowcast value is *R*_*t*1,RKI_ = 0.82 [47]). Similarly, we estimated *R*_*t*2_ = 0.95 (*R*_*t*2,RKI_ = 1.1) over the second window from June 9 to June 14 after several NPIs were lifted. Economic activity was measured at the industry level (NACE Rev. 2) based on the ifo Business Survey, a monthly survey among German firm managers that includes approximately 9,000 responses with respect to their current and expected business activities (see [48] for details on survey methodology).

The second step in estimating the economic consequences of different NPI scenarios, which vary in their degree of stringency, is to simulate the economic activity associated with each scenario. All simulations start during the initial lockdown phase when *R*_*t*1_ = 0.84 and then, for each set of contact rates considered, proceed forward in time until either the modelled number of newly reported cases falls below a threshold *I*_max_, or a maximum time threshold *T*_*M*_ = 300 d is reached. The threshold *I*_max_ depends on the capacity of the public health system to control the epidemic through contact tracing and isolation. For Germany, this threshold was estimated early in the pandemic to be 300 newly reported cases per day, based on the number of health departments (see [28] for details). The length of time from shutdown initiation until one of the thresholds *I*_max_ or *T*_*M*_ is reached, *T*_*S*_, defines the duration of lockdown, over which the economic costs are calculated.

Each scenario is defined by time-dependent contact rates *β*_*j*_. For each set of *β*_*j*_, the behavioural-epidemiological model is simulated to obtain the lockdown duration and corresponding *R*_*t*_ values throughout the lockdown. Given the *R*_*t*_ values, the sector-specific economic activity can then be quantified throughout the lockdown via the regression estimated in step one above (see Annex C for full details). The total cost for scenario *i* is then obtained by summing over the lockdown period as

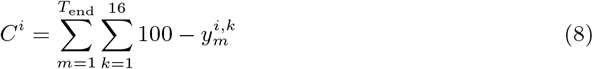

where 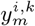 is the level of economic activity of sector *k* compared to pre-shutdown levels in scenario *i* and month *m*. Full details of how 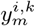 are calculated over the lockdown periods can be found in Annex C.

We then express the total cost for each scenario relative to a reference scenario in which *R*_*t*2_ = *R*_*t*1_ and then plot total relative cost *C*^*i*^ vs. 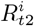. First, this allows us to visualise how the economic impact of a given NPI scenario, expressed as total relative cost, is driven by the stringency of lockdown measures, measured as *R*_*t*_. Second, this approach allows us to compare different NPI strategies based on where they fall along the relative cost curve.

#### Scenario transfer

To transfer the NPI strategy of a reference country to the focal country, we first identify the dates of NPI changes in both countries. Next, the model parameters are estimated for both countries separately via model fit to each country’s COVID-19 incidence data. For the scenario run, we use the first NPI dates and associated disease transmission rates of the focal country, until the epidemic wave has fully evolved (i.e. the first two NPI dates of the focal country are fixed). The subsequent NPI dates and disease transmission rates are then transferred from the reference country to simulate how the wave would have further developed in the focal country under the reference country’s NPI strategy. To transfer contact rates between nations, we assume that NPIs have the same proportional effect in each nation, but NFCs cause the proportionality constants to differ between countries. We therefore calculate the relativised, transferred contact rates *τ* ^*T*^ from the reference country (R) to the focal country (F) as:

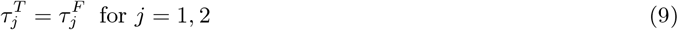

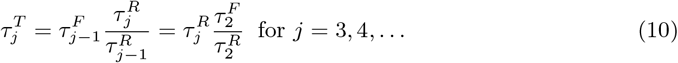

The relativised contact rate transfers the effects of NPIs *per se*, but does not account for differences in autonomous behavioural responses between reference and focal nations. To account for behavioural differences, the estimated risk aversion parameter, Δ*v*, can also be transferred from reference to the focal nation. To help separate these effects, we consider scenarios with only NPI transfer, and with both NPI and risk aversion transfer.

## Results

The dynamics of the first COVID-19 wave in DE can be successfully reproduced with the coupled epidemiological (Eqs. (1) - (3)) and behavioural (Eq.(7)) model (Fig. 3A). Only the small summer peak could not be captured by the model, as it cannot be explained by NPIs. Similarly, the incidence curve for NZ can also be adequately reproduced (Fig. 3B), even though the initial increase in cases is so steep that the model cannot keep up and therefore creates a more gradual increase that, compared to the data, starts slightly earlier. The coupled model can describe the CH data as well, where there was a rapid initial increase in cases with a gradual decline thereafter (Fig. 3C). Overall, the epidemiological development during the first wave is qualitatively similar in DE, CH, and NZ. In all three cases, we were able to unambiguously identify reasonable estimates for all model parameters.

**Fig 3.**
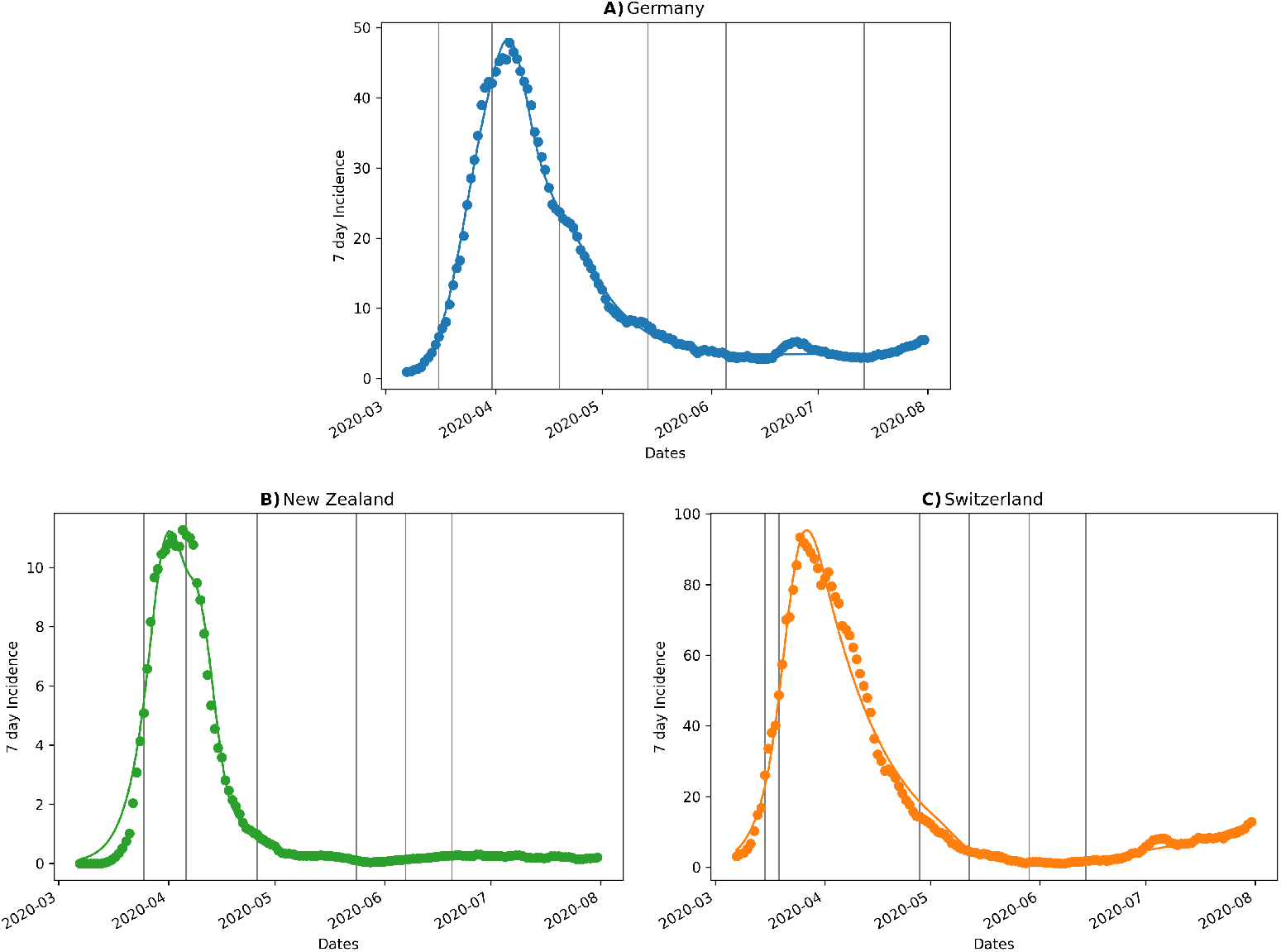
Fit of the coupled behavioural and epidemiological model to incidence data from A) Germany (upper panel), B) New Zealand (lower left), and C) Switzerland (lower right) during the first wave in Spring 2020.

Transferring the relativised transmission rate (Eq. (9)) from NZ to DE from the second NPI period onward demonstrates that the first wave would have been stopped earlier than was actually observed in DE (Fig. 4A,C). Specifically, combining the NZ relativised transmission rate with DE’s level of risk aversion would have led to the outbreak ending in early May 2020 (Fig. 4A), while the more conservative level of risk aversion apparent in NZ would have helped end the outbreak by late April (Fig. 4C).

**Fig 4.**
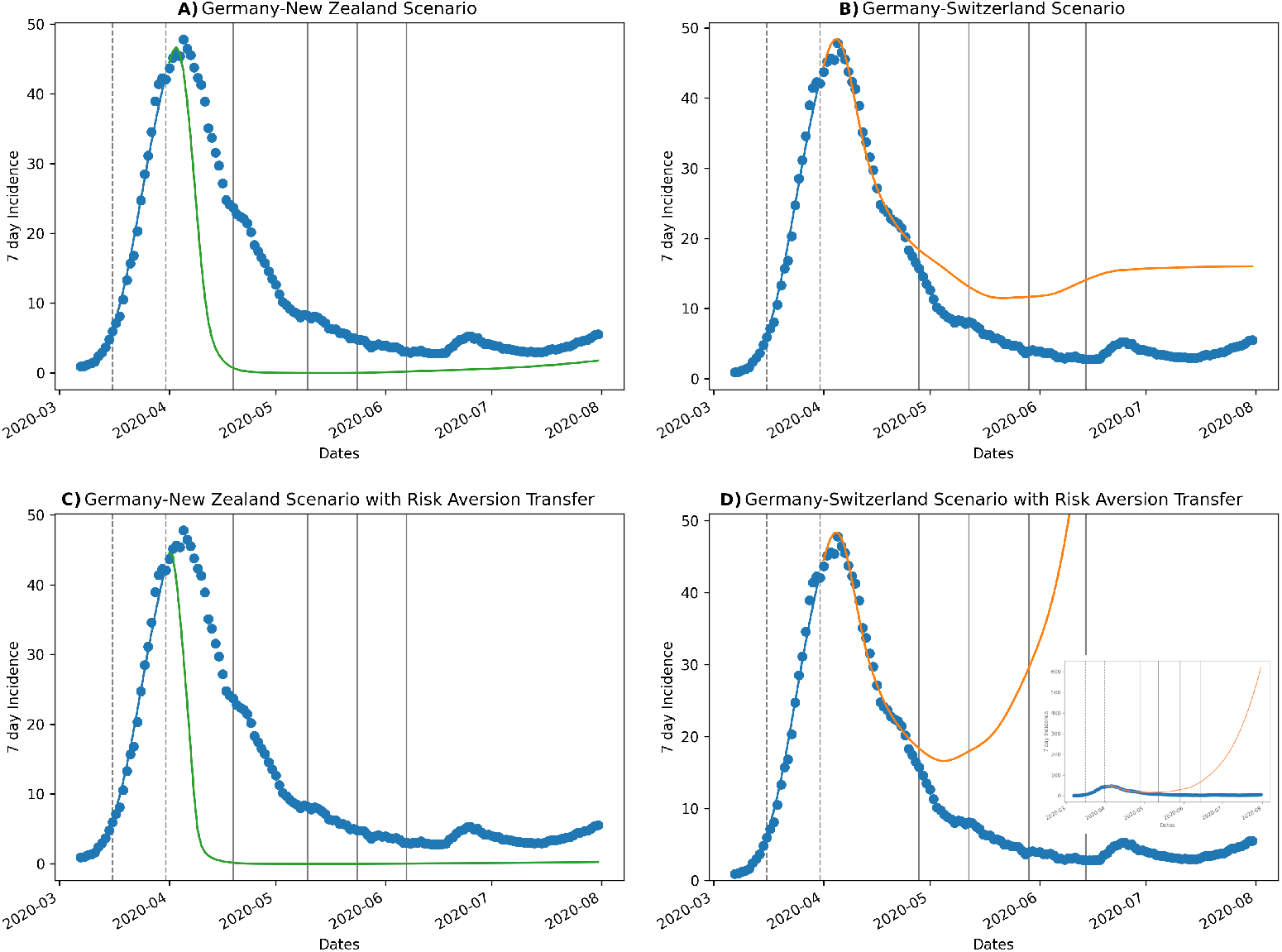
A) New Zealand (upper left panel) and B) Switzerland (upper right panel) scenarios transferred to Germany from the second NPI date (second vertical dashed line) onward. The lower panels show the effect of also transferring the estimated risk aversion from the reference countries C) New Zealand and D) Switzerland to Germany.

Applying the CH scenario to DE (Fig. 4B,D), we see that the first NPIs seem to be enough to break the wave. In the transfer scenario with DE risk aversion (Fig. 4B), the subsequent NPIs appear to be only stringent enough to create a dynamic balance with the 7d incidence staying around 15 d^*−*1^ for about 3 months. In stark contrast, the transfer scenario with CH risk aversion, which was markedly lower than for DE, would have resulted in a very large second wave during summer 2020 (Fig. 4D). Comparing panels B and D in Fig. 4 shows that the autonomous behavioural response, which is governed by risk aversion, can qualitatively change the outcome of a given NPI regime when pronounced differences in risk aversion occur.

The parameterised models also contain information on reductions in social contacts via the utility function (Eq. (6)). Specifically, the utility function quantifies the reduction in contacts relative to pre-pandemic levels. The actual DE NPIs and the DE risk aversion reduce contacts to a little less than 60% of pre-pandemic levels (Fig. 5, blue curve), and the CH scenario with DE risk aversion also causes a similar reduction in contacts (Fig. 5, orange solid curve). In contrast, the NZ scenario with DE risk aversion immediately reduces relative contacts to 30% from the onset of the first wave, which then gradually increases to values above the DE and CH scenarios, finally decreasing again to similar values as for DE and CH during the summer of 2020 (Fig. 5, green solid curve). Transferring the NZ risk aversion results in no important difference (Fig. 5, green dashed curve). The explosive increase in cases in the CH scenario with transferred risk aversion ultimately breaks the assumption of the behavioural model that the infection numbers are small, which results in relative contacts becoming negative during summer 2020 (Fig. 5 orange dashed curve). However, we have included this result (truncated at zero) both for the sake of completeness, and because it offers qualitative insight. Specifically, it seems reasonable that the large second wave would indeed drive contacts below the level observed during the first wave due to the autonomous behavioural response. As an external check on our coupled model, we note that the relative contact reduction in DE estimated from our model (Fig. 5, blue curve) agrees remarkably well with the results presented by Rüdiger et al. 2021 ( [49], Fig. 2B). Importantly, they used mobile phone data to estimate a contact reduction of approximately 50%, whereas our estimates were based entirely on incidence data. The concordance between these differently estimated results suggests both that our behavioural submodel is capturing the key drivers of contact reduction, and is estimable from incidence data alone.

**Fig 5.**
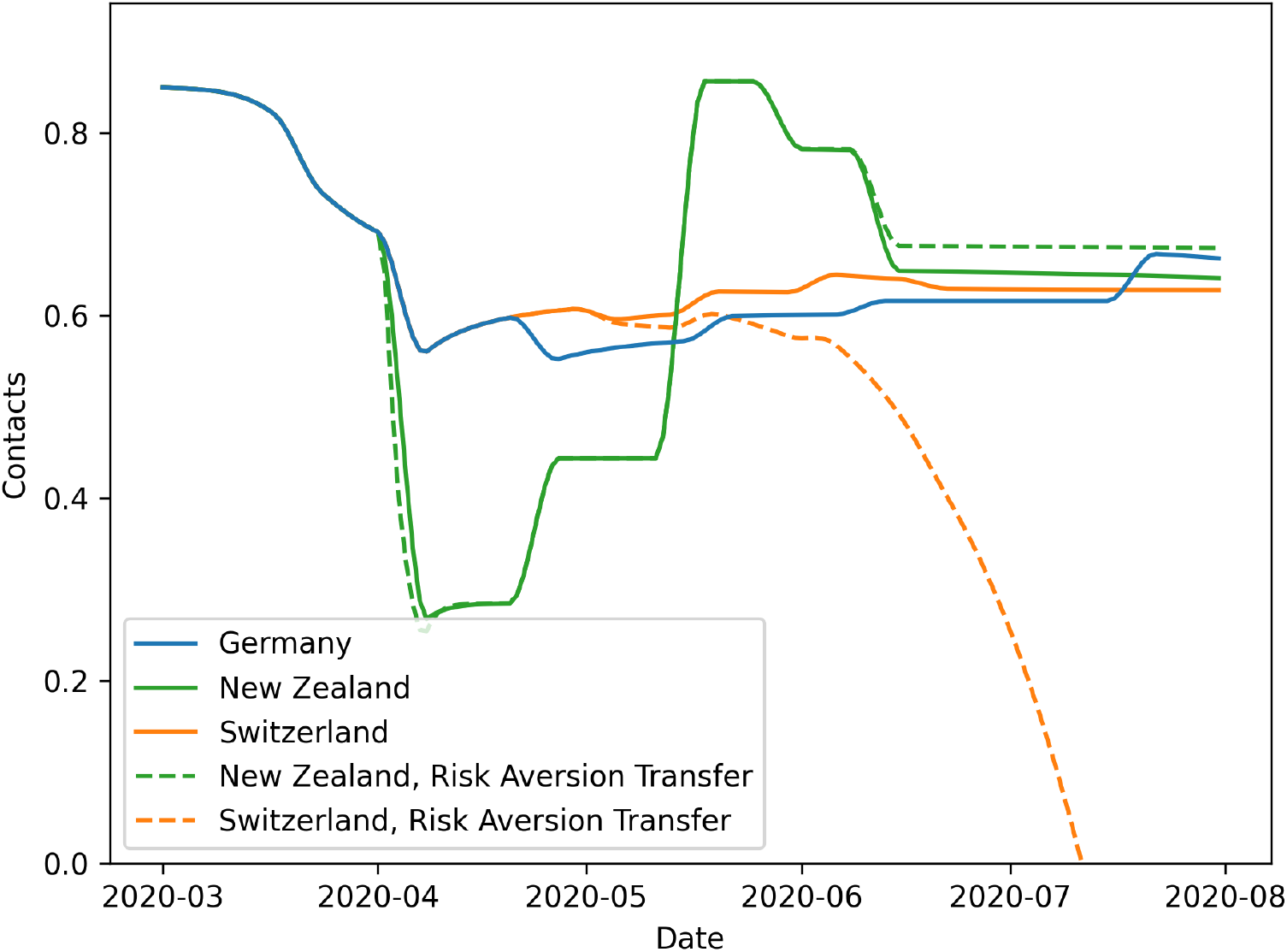
The dynamics of relative contact reduction for Germany and for the New Zealand and Switzerland transfer scenarios with and without risk aversion transfer.

The relative cost function from our economic model is convex and asymmetric, showing that long-run costs increase faster and reach higher levels under very lenient NPI scenarios than under stricter NPI scenarios (Fig. 6). Economic activity is more sensitive to long shutdown duration (lenient NPIs) than to shorter but stricter shutdowns. Germany imposed measures that resulted in somewhat higher than optimal economic costs (Fig. 6, blue point). The minimum is found at the midpoint between the shutdown costs (*R*_*t*1_ = *R*_*t*2_ = 0.84) and partial opening costs (*R*_*t*2_ = 0.95), suggesting that economic costs would have been minimised if the partial opening measures in DE had been slightly more lenient compared to the actually implemented strategy.

**Fig 6.**
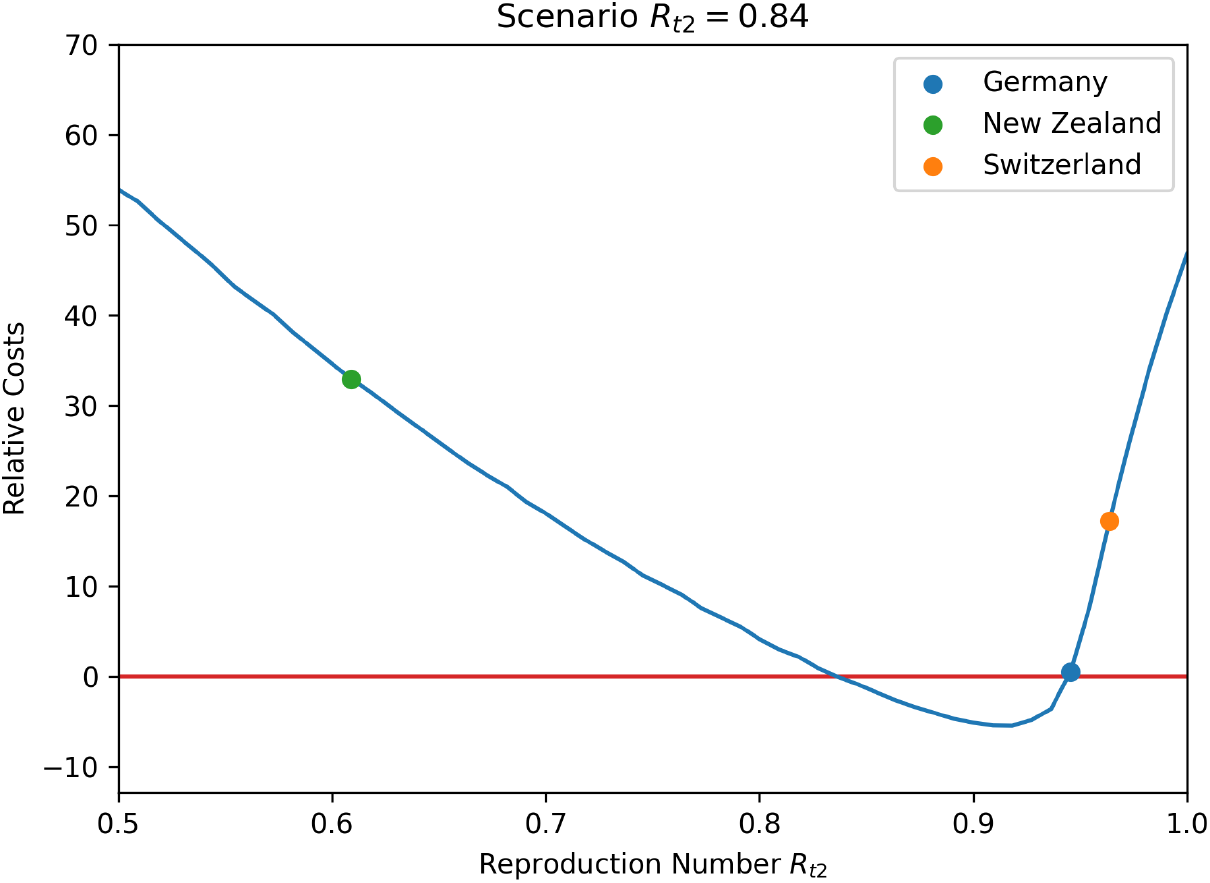
Relative economic costs of the three different NPI strategies in the context of Germany.

**Fig 7.**
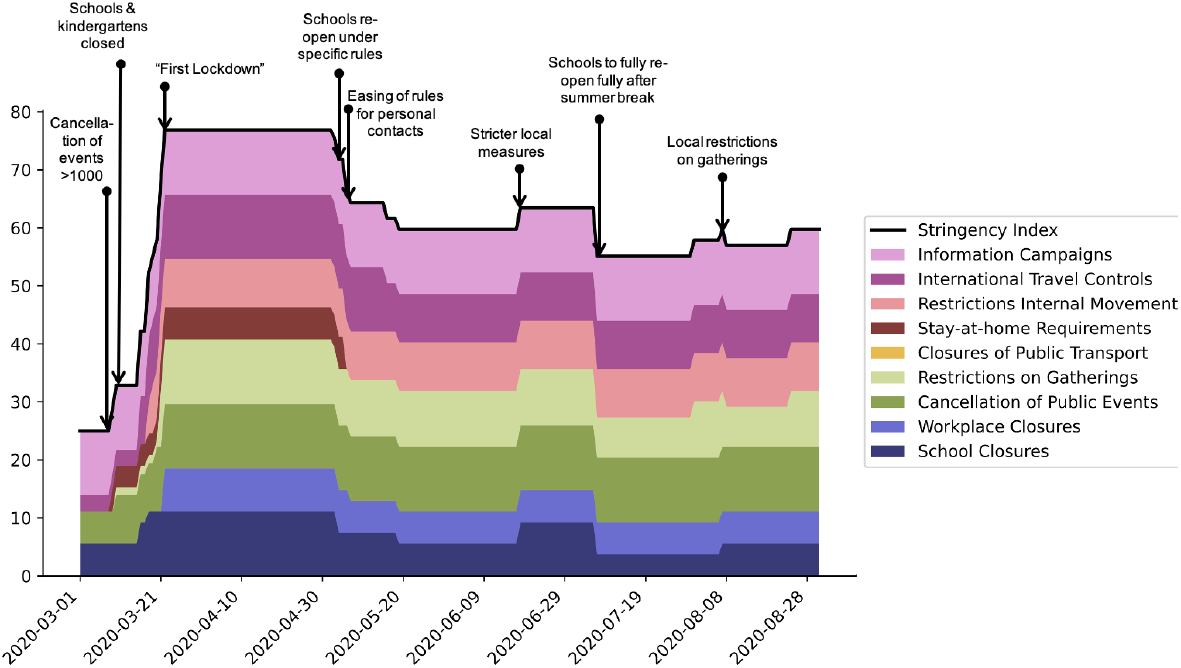
Stringency index composition: Germany (Mar-Aug 2020)

**Fig 8.**
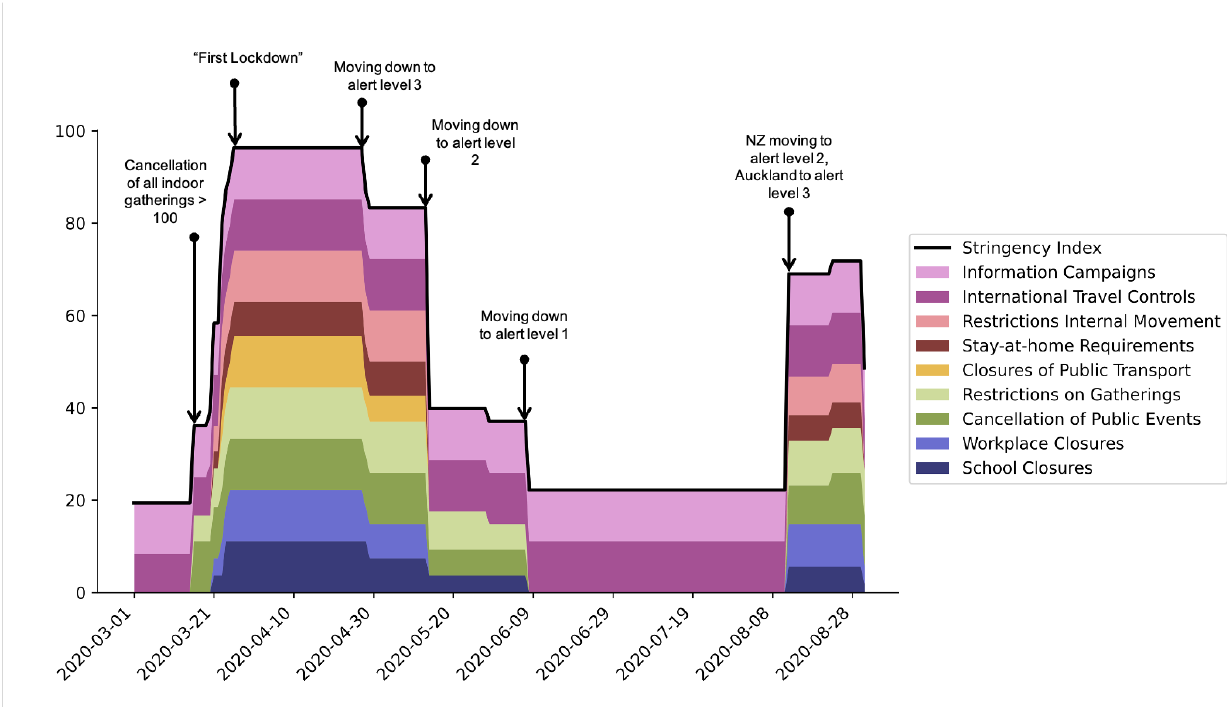
Stringency index composition: New Zealand (Mar-Aug 2020)

**Fig 9.**
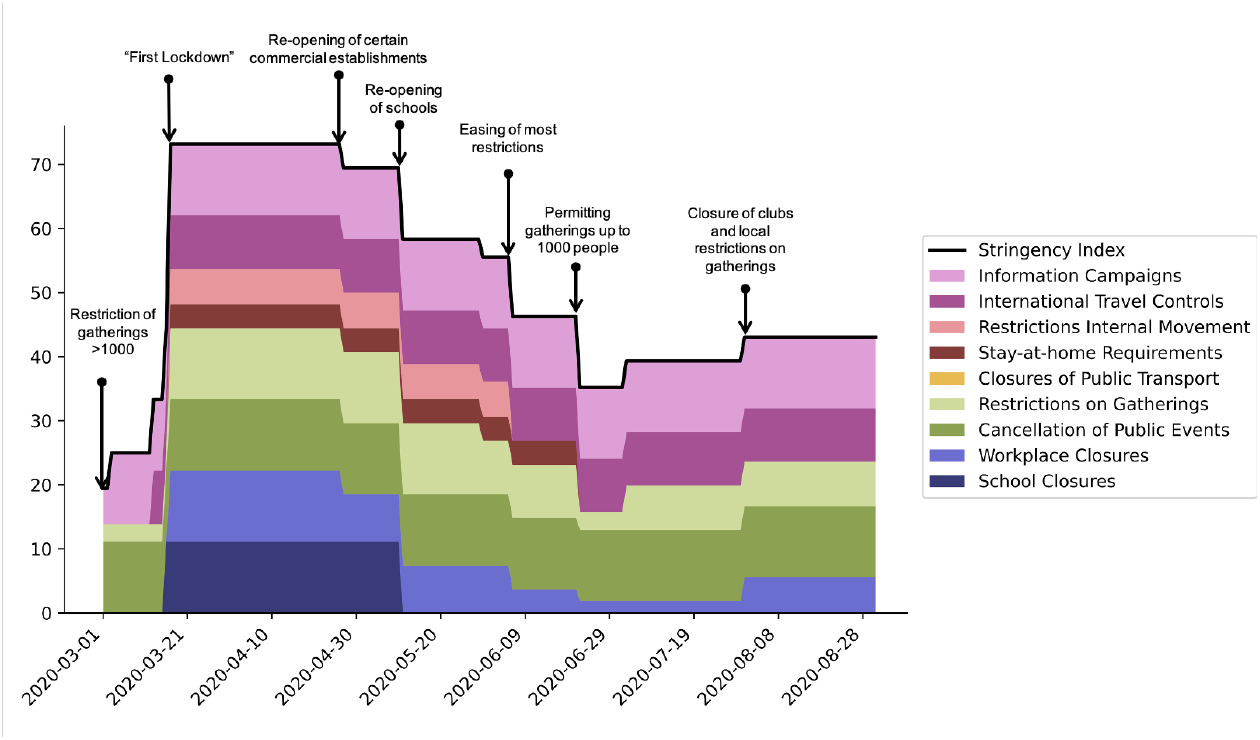
Stringency index composition: Switzerland (Mar-Aug 2020)

Despite resulting in divergent epidemiological outcomes (Fig. 4), and similarly large differences in social outcomes (Fig. 5), the economic model suggests that both the NZ and CH scenarios, when viewed in the context of DE, were suboptimal and would have resulted in substantial relative cost increases (Fig. 6). Here, we focus only on the cases where German risk aversion was used in both the CH and NZ scenarios, as: 1) differences in risk aversion had negligible economic consequences in the NZ scenarios, and 2) CH risk aversion resulted in *R*_*t*2_ *>* 1, which violates one of the economic model’s assumptions. In the case of NZ, the more stringent NPI response quickly brings the first wave to heel, but results in substantially increased short-term economic impacts (Fig. 6, orange point). In contrast, the more lenient CH strategy has lower initial economic impact, but the resulting prolonged outbreak causes costs to accumulate over the longer term, yielding an even higher relative cost increase than for the NZ scenario (Fig. 6, green point).

## Discussion

Being better prepared for the next pandemic requires that we learn as much as possible from the unprecedented volume of data generated by COVID-19. Cross-national comparative studies are foundational components of this learning process, but require special care given the myriad differences among nations (i.e., NFCs) that may contribute to broadly different outcomes. In the case of NPI strategies, such comparisons are further complicated by NPIs being applied as packages of interventions, which makes it more difficult to quantify the effects of individual measures in isolation. Conventional approaches to inter-country comparisons focus on teasing apart individual effects of interventions and ranking them in terms of average effect magnitude [1, 5]. Finally, most assessments of NPI effectiveness focus solely on epidemiological outcomes, neglecting social and economic dimensions. In contrast, we have taken a novel, scenario-based approach that can control for differences in NFCs, recognises the non-independence of NPIs as applied in practice, and explicitly considers the economic and social consequences of NPIs in the context of a focal country.

Choosing DE as our focal nation, and the first COVID-19 wave in Spring 2020 as our time horizon, we have considered two scenarios with contrasting properties. New Zealand implemented a very strict NPI strategy focused around an intense national lockdown, while CH choose a much more lenient strategy specifically to avoid the economic and social consequences a hard nationwide lockdown entails. When transferred to the context of DE, these strategies produce markedly different epidemiological outcomes both relative to each other and relative to DE’s realised NPI strategy. The NZ strategy results in a sharp, immediate decline in cases and near total elimination of the outbreak in a very short time. When the NZ NPI strategy is accompanied by the NZ level of risk aversion, which was higher than in DE, the outbreak is extinguished a couple of weeks sooner, but the qualitative pattern does not change.

Switzerland’s strategy, coupled with DE risk aversion, stops further increases in cases, but also fails to drive incidence levels towards zero. In contrast to the NZ scenarios, accounting for the lower level of risk aversion in CH compared to DE changes the outcome of the simulated transfer scenario both quantitatively and qualitatively. Specifically, a clear and much larger second wave appears following the first, indicating that the later-stage CH NPIs would have been inadequate to prevent a further outbreak in the context of DE. This qualitative shift occurs because the CH NPIs were at the limit of being able to contain a further outbreak in the DE context. Thus, a change in average individual risk aversion was sufficient to tip that fragile balance in direction of another surge of infections.

The different strategies employed by DE, NZ, and CH have substantially varied social costs. It is immediately apparent that the NZ strategy, when transferred to DE, would have resulted in dramatic reductions in social contacts, either with or without NZ risk aversion. Given the strong social and political reactions to lockdowns and loss of contacts that were actually observed in Germany, such further reductions could have been extremely disruptive. In the CH transfer cases, the difference in social costs, driven by differences between DE and CH levels of risk aversion, is striking. In the DE risk aversion case, the predicted reduction in contacts is comparable to what was actually observed in Germany, and much less than either of the NZ transfer scenarios, at least until mid-May 2020. However, in the CH transfer scenario with CH risk aversion, contacts are quickly driven to zero, resulting from a somewhat counter-intuitive feedback loop. Specifically, lower risk aversion interacts with weaker NPIs to produce much higher case counts. The exploding case numbers then drive an almost total loss of social contacts due to individuals’ autonomous behavioural responses, even though risk aversion was markedly lower in CH than in DE. In other words, a severe enough outbreak can trigger dramatic reductions in social contacts even when risk aversion is low (but non-zero). However, the behavioural model assumes low infection numbers. Therefore, at least the quantitative results of the reduction in social contacts should be taken with caution. This is caveat is discussed in more detail below.

When viewed through the lens of our DE-specific economic model, both the NZ and CH scenarios prove to substantially increase relative costs, but for contrasting reasons. Specifically, the NZ scenario incurs the immediate costs of a very strict lockdown, while only yielding modest epidemiological gains. In contrast, CH’s more relaxed approach gradually accumulates costs resulting from a sustained, low-intensity lockdown and concomitant reductions in business activity and efficiency. Relative to the ‘middle road’ strategy DE chose to implement, both of these reference approaches perform poorly from an economic perspective, while only NZ’s approach would have yielded epidemiological gains in the simulated DE national framework. Though we were not able to calculate relative economic costs in the CH scenario with CH risk aversion due to a violation of the economic model’s assumptions, we can surely conclude the relative costs would have been even higher because it would have taken longer to bring the daily number of cases back down to the threshold *I*_max_.

Our goals here were to develop an elemental approach to scenario-based, inter-country comparisons of NPI strategies, and to demonstrate the insights that can be gained from this framework. We have therefore kept our component models as simple as possible. A clear limitation of this approach is the fact that, in its current state, the economic model can’t handle scenarios that feature *R*_*t*2_ *>* 1, as happened in the CH scenario with CH risk aversion. This scenario also highlights a limitation of the behavioural model, which is the assumption of small case numbers *I*. This becomes problematic if we try to calculate the relative contact reduction from Eq. (6) (see Fig. 5, orange dashed curve). In the asymptotic case for *I → ∞*, the epidemiological part of the coupled models (1) - (3) and (7) would also break down. However, large *I* values are only gradually reached from June 2020 onward and the risk aversion parameter in Eq. (7) is generally small compared to the cases *I*, with Δ*v <* 10^*−*4^. This leads to a situation, where the equations stay stable until the point where the threshold 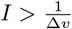 is reached. For the CH scenario with CH risk aversion, this stability threshold is not exceeded. One could, of course, envision more elaborate versions of any of the pieces of our coupled model. Such embellishments would need to carefully balance realism against tractability. In particular, our approach hinges on being able to reliably estimate model parameters from epidemiological and economic data. While more detailed models might be easier to justify mechanistically, this may come at the cost of parameter identifiability, which is an issue that frequently plagues complex compartment-type disease models [50, 51]. Despite these concerns, we believe that significant opportunities exist to build on the framework we have established here.

Finally, it is important to realise that the conclusions drawn from our scenario-based analyses are specific to a particular focal country, in this case DE. Our approach is silent with respect to the optimality of the reference country’s strategy *in the context of the reference country*. For example, we concluded that DE’s realised NPI strategy achieved a more ideal balance between epidemiological and economic goals than NZ’s strategy would have, had it been implemented in DE. However, we draw no conclusions about how good NZ’s strategy was for NZ. Similarly, we also did not explore how well DE’s strategy would have performed in either NZ or CH. Doing so would require parameterising our economic model for these countries, and then treating each as a focal country as we’ve done here for DE. Indeed, expanding the catalogue of both focal and reference nations to which our approach can be applied is a clear priority for subsequent work. We are therefore currently compiling the global databases that would support more extensive comparisons, and intend to explore a broader range of case studies in the future.

## Annex A: Stringency index breakdown and timeline

## Annex B: Derivation of reduced-form behavioural model

The behavioral model (6) is a reduced form version of the model developed in [27], and closely related to the approach proposed in [52]. According to that model, the expected present value of life-time utility of a susceptible individual at time *t* is given by

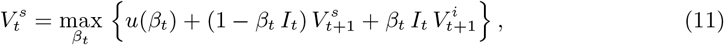

where subscripts *t* denote the current time period, *t* + 1 the next time period, etc. With probability 1 *− β*_*t*_ *I*_*t*_ the individual stays susceptible and then will enjoy the expected present value life-time utility of a susceptible individual in the next period, 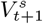. With probability *β*_*t*_ *I*_*t*_, the individual will get infected and the expected present value life-time utility is reduced to that of an infected individual, 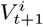, which may in the subsequent period stay infected, recover, or die. In a fully dynamic and recursive model, all these present value utilities need to be derived [27, 52]. Here, we assume that the present value utilities are independent of time, such that we can drop the time index of *V* ^*i*^ and *V* ^*s*^, and (11) becomes

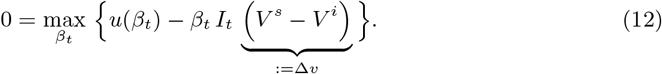

The zero on the left-hand side is a matter of normalisation of the utility function. The relevant part is the model of behaviour, i.e. the maximisation of the right-hand side of (12), which is (7). The corresponding first-order condition reads *u*^*′*^(*β*) = *I* Δ*v*. Using the specification (5) of the utility function, this becomes

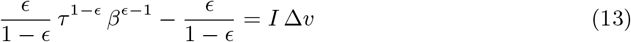

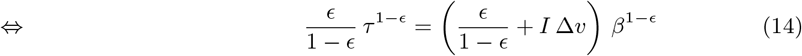

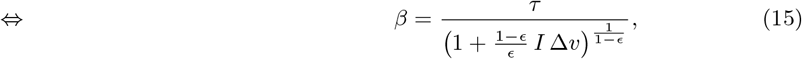

which is equation (7).

## Annex C: Economic model details and parameters

Here, we provide further details of how the sector-specific economic costs are calculated for a given scenario over the resulting lockdown duration. Following Dorn et al. (2022) [28] we calculate the activity of each sector for six different phases related to the shutdown: (a) pre-shutdown economic activity, normalised to 1, (b) shutdown, (c) initial transition period after implementation of policy change, (d) full transition period under partial restrictions, (e) recovery period, and (f) complete recovery. The expected duration of recovery Δ*t*_3_ was inferred from a special question in the ifo Business survey in June 2020. Given the regression relationship between *R*_*t*_ and 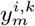, economic costs for each phase are calculated as

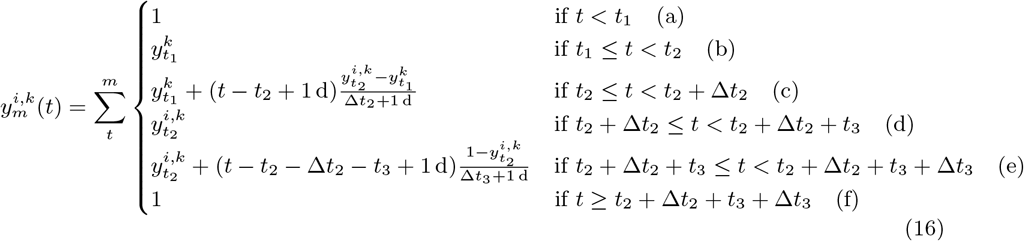

where the dates *t*_1_, *t*_2_, *t*_3_ denote the shut-down, the partial re-opening, and the start of the reopening phase. The durations Δ*t*_2_, Δ*t*_3_ denote the initial adjustment time of the re-opening and the time of the recovery. 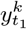 is the economic activity of sector *k* during shutdown (*t*_1_ until *t*_2_) and 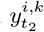 is the economic activity of sector *k* after the first re-opening under scenario *i* (*t*_2_ + Δ*t*_2_ until *t*_2_ + Δ*t*_2_ + *t*_3_). The economic costs are aggregated over the sectors, weighted by their shares out of overall gross value added, to monthly values, indicated by. 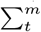.

**Table 1.** Model summary.

| Parameter | Value (baseline) | Interpretation | Source |
| --- | --- | --- | --- |
| $R_{t1}$ | 0.84 | Stringency of shutdown | model |
| $R_{t2}$ | 0.95 | Stringency of policy change | model |
| $y_{t_1}^k$ | [30, ..., 75] | Economic activity of sector $k$ under shutdown | ifo Business Survey |
| $y_{t_2}^{i,k}$ | [45, ..., 90] | Economic activity of sector $k$ after first openings under scenario $i$ | ifo Business Survey |
| $T_{\text{end}}$ | 36 mo | Time horizon of simulation | Exogenous |
| $T_M$ | 300 d | Threshold duration for reopening | Estimate |
| $I_{\text{max}}$ | 300 d<sup>-1</sup> | Threshold cases for reopening | Estimate |
| $t_1$ | 2020-03-19 | Calendar day of shut-down implementation | Government announcement |
| $t_2$ | 2020-04-20 | Calendar day of policy change | Government announcement |
| $t_3$ | 60 d | Start of re-opening phase | Exogenous |
| $\Delta t_2$ | 21 d | Initial adjustment duration | Exogenous |
| $\Delta t_3^{i,k}$ | 8.2 mo (Min., J)<br>10.5 mo (Max., I)<br>9.0 mo (Mean) | Duration of recovery under scenario $i$ in sector $k$ | ifo Business Survey |

## Data Availability

All data used in the manuscript are available online at https://rodare.hzdr.de/record/2467

